# Predicting 30-Day Mortality and Readmission Using Hospital Discharge Summaries: A Comparative Analysis of Machine Learning Models, Large Language Models, and Physicians

**DOI:** 10.1101/2025.03.26.25324714

**Authors:** Alexander Scarlat, Francis X. Campion

## Abstract

This study evaluates the comparative performance of trained machine learning models, commercial off-the-shelf (COTS) large language models (LLMs), and physicians in predicting 30-day mortality and readmission using discharge summaries from the MIMIC-IV dataset. While machine learning models demonstrated superior performance compared to both LLMs and human evaluators, none of the approaches achieved clinical-grade reliability, defined by us as a Matthews Correlation Coefficient > 0.8. Interestingly, all methods performed better at predicting mortality (3% incidence) than readmission (21.1% incidence), suggesting that the signal-to-noise ratio may be higher when predicting mortality. The universal difficulty both machines and humans encountered in these prediction tasks indicates fundamental challenges in forecasting post-discharge outcomes from discharge summaries alone. These findings highlight both the potential and current limitations of artificial intelligence in clinical prediction tasks, while emphasizing the inherent complexity of such predictions regardless of the approach used.

## Introduction

Accurate prediction of 30-day mortality and readmission risk is crucial for clinical assessment and resource optimization [1, 2]. Discharge summaries, which contain detailed information of patient hospital stays and care plans, represent a rich but challenging source for predictive modeling. While traditional statistical methods have shown promise, the emergence of machine learning and natural language processing has opened new possibilities for analyzing these complex clinical narratives. Historically, artificial intelligence (AI) methods for predicting readmission have used machine learning models trained on discrete data such as health insurance claims data or clinical data such as diagnosis codes, laboratory values, and vital signs from the electronic health record [3, 4]; however, implementing such solutions is challenging and usually limited by data constraints of individual hospitals. In 2020, Rumshisky and colleagues incorporated natural language processing on electronic health record (EHR) text, combined with machine learning of discrete data to outperform human raters in predicting psychiatric readmissions, revealing the nuanced capabilities of AI in processing narrative summaries [5].

Recent studies have demonstrated the potential of more advanced machine learning, and new AI applications using Large Language Models (LLMs) to evaluate EHR text documents. Innovative open-source projects such as Meditron-70B from researchers at Yale University are promising. They have demonstrated the use of extended pre-training using a medical document corpus and fine tuning to improve performance of Llama-2 for medical question and answering tasks but have not yet published on prediction tasks [6].

However, the clinical applicability of these technologies requires thorough validation, particularly in comparison to human expert performance. In particular, clinicians may be attracted to use general purpose LLMs because of their ease-of-use, but risks related to bias, privacy, and hallucination have the potential to cause patient harm [7]. The use of general purpose LLMs for consequential classification and prediction tasks such as hospital readmission and mortality remain to be critically evaluated. In this study, we demonstrate the use of a standardized test data set of EHR discharge summaries to compare the tasks of prediction of 30-day readmission and 30-day mortality after hospitalization for adult patients. We compare performance of machine learning, neural networks, and general purpose LLMs with physician performance.

## Methods

### Data Source and Cohort Selection

We used data from the Medical Information Mart for Intensive Care IV (MIMIC-IV) database, which contains 331,794 discharge summaries corresponding to 145,915 unique individuals [8]. Downstream embedding models accepted a limited maximum number of tokens, requiring the exclusion of all discharge summaries longer than 8,190 tokens. This requirement removed approximately 0.18% of available samples. We also excluded individuals who died during the index hospital admission (2% of the dataset). This yielded 323,076 discharge summaries from 141,987 unique individuals, of which the average discharge summary contained 1,672 words (99% of summaries remain under 3,987 words). In this overall cohort, mortality within 30 days of discharge was 2.94% and the 30-day readmission rate was 20.93%. From this pool of 323,076 eligible discharge summaries, we randomly selected 20,000 summaries for further analysis. These 20,000 summaries corresponded to 17,572 unique individuals. Within this 20,000-sample subset, the 30-day mortality rate was 3.0%, and the 30-day readmission rate was 21.1%.

### Train/Test Split by Individuals

To avoid data leakage, we split the dataset at the level of the patient rather than the discharge (Figure 1). Splitting by discharges would risk placing different samples from the same patient into both training and testing sets. We therefore assigned 80% of individuals to training (n = 14,057 individuals; 16,013 discharges) and 20% of individuals to testing (n = 3,515 individuals; 3,987 discharges). An individual may appear in the train or in the test subset, but not in both.

**Figure 1.**
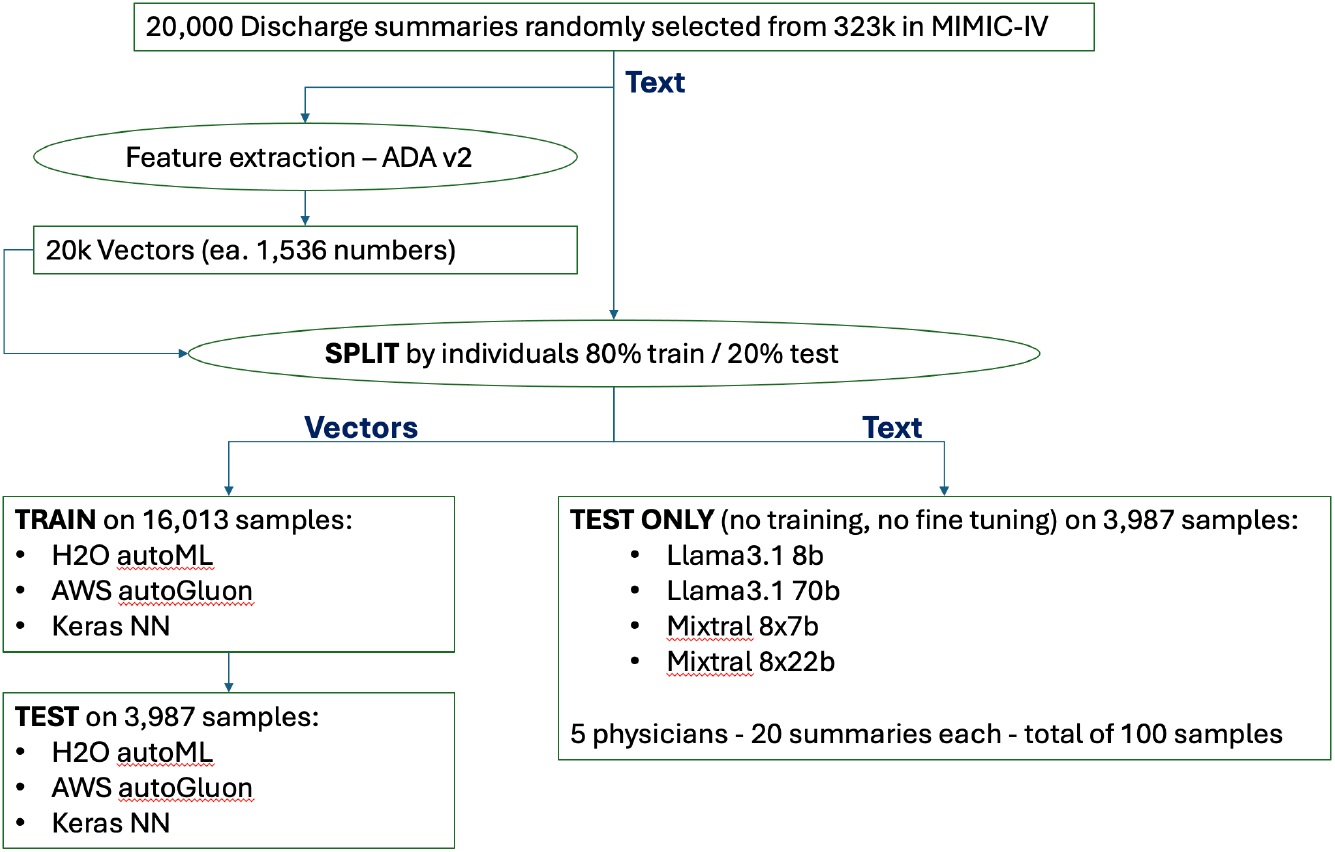
Data Selection, Preprocessing, and Modeling Pipeline.

### Evaluation Metric: Matthews Correlation Coefficient (MCC)

We selected the Matthews Correlation Coefficient (MCC) as our primary evaluation metric, utilizing all four components of the confusion matrix—true positives, false positives, true negatives, and false negatives (TP, FP, TN, FN). The MCC ranges from −1 to +1: +1 indicates perfect agreement between predictions and ground truth, 0 indicates random (or chance-level) performance, and −1 indicates complete disagreement (i.e., inverse prediction).

This makes MCC especially suitable for our imbalanced dataset, as it does not overly reward correct predictions on the majority class or penalize misclassifications of the minority class. In contrast, common metrics such as the F1 score (which ignores TN) and the area under the ROC curve (which ignores both TN and FN) can be misleading when class distributions are heavily skewed. By considering each type of prediction error and success, MCC provides a more comprehensive and balanced assessment of model performance [9].

### Baselines

We established three simple baseline methods to compare the performance of each model:

1. Human benchmark: Five physicians each independently evaluated 20 randomly selected discharge summaries (100 in total) from the test subset, providing two binary predictions for each case: (1) whether the patient died within 30 days of discharge and (2) whether the patient was readmitted during that interval.
2. Majority Guessing: For each of the two binary outcomes (30-day mortality and 30-day readmission), we predicted the majority class. For instance, if 97% of patients in the test set survived past 30 days, we predicted survival for all patients; similarly, if 80% of patients were not readmitted, we predicted no readmissions for all patients.
3. Ground-Truth Shuffling: Using the same 3,987 test subset, we shuffled the outcome labels (while preserving overall event rates) to produce another naive prediction baseline. This approach maintains the original proportion of positive and negative events but eliminates any relationship between the input features and the actual outcomes.

### Feature Extraction (Embedding)

To transform the free-text discharge summaries into a numerical representation suitable for downstream modeling, we generated 1,536-dimensional embedding vectors using the ADA v2 model (“ada002”) from OpenAI, with no external exposure of data. This embedding step converts each discharge summary into a fixed-length numeric vector reflecting its semantic content. After embedding, we obtained 16,013 vectors for the training set and 3,987 vectors for the testing set, each vector comprising a list of 1,536 numbers. These numbers are the features extracted from the text summarizing its semantic meaning.

### Downstream Predictive Modeling

Using the resulting 1,536-dimensional vector embeddings, we trained three supervised learning models on the training set: a Keras-based neural network, the autoML from H2O, and AWS AutoGluon frameworks. All models received the 16,013 vectors (each representing one discharge summary) as inputs. We then evaluated model performance on the reserved test set (3,987 vectorized discharge summaries).

### LLM Zero Shot Evaluation

Additionally, we evaluated zero-shot performance of large language models (LLMs): Llama3.1 8B and 70B, Mixtral 8×7B and 8×22B directly on the text of the same 3,987-test-sample set, without performing any embedding or fine-tuning. The prompts used for each LLM were as follows:

#### Mortality prompt

*“Read the discharge summary and then predict whether the patient will die within 30 days of discharge as 1, and if the patient lives as 0*.*”*

#### Readmission prompt

*“Read the discharge summary and then predict whether the patient is readmitted within 30 days of discharge as 1, and if the patient is not readmitted as 0*.*”*

## Results

| Mortality - MCC results |  |
| --- | --- |
| Experiment | MCC |
| Majority guessing | 0 |
| Shuffle the ground truth guess | 0.023 |
| Llama3-8B | 0.111 |
| *Physicians* | 0.208 |
| Mixtral 8x7b | 0.255 |
| Llama3-70B | 0.271 |
| Mixtral 8x22b | 0.358 |
| AWS AutoGluon trained on 16k | 0.458 |
| H2O AutoML trained on 16k | 0.484 |
| Keras NN trained on 16k | **0.491** |

Readmission - MCC results
| Experiment | MCC |
| --- | --- |
| Majority guessing | 0 |
| Shuffle the ground truth guess | 0.026 |
| Llama3-8B | 0.028 |
| Mixtral 8x7b | 0.079 |
| Llama3-70B | 0.103 |
| Mixtral 8x22b | 0.153 |
| *Physicians* | *0.154* |
| H2O AutoML trained on 16k | 0.21 |
| AWS AutoGluon trained on 16k | 0.212 |
| Keras NN trained on 16k | **0.213** |

**Note:** All MCC results come from the same 3,987 test subset, except the physician results, which are based on only 100 discharge summaries randomly selected from this subset.

## Discussion

This study highlights the complex challenges associated with utilizing discharge summaries to predict 30-day mortality and readmission rates. A particularly notable finding is the universal difficulty encountered by both machine learning models and human experts in achieving highly reliable predictions. Neither advanced AI systems nor experienced physicians met our threshold for reliability, defined as an MCC greater than 0.8. This underscores the inherent complexity of predicting post-discharge events using narrative summaries alone and raises questions about the adequacy of such summaries as standalone predictive tools.

Supervised machine learning models consistently outperformed COTS LLMs and human evaluators, demonstrating the value of task-specific training on domain-relevant datasets for clinical prediction tasks. However, the inability of any model to reach the desired reliability threshold indicates that current ML technologies remain insufficient for autonomous application in these specific clinical decision-making contexts.

An unexpected yet significant observation was the consistently superior performance in predicting mortality (3% incidence) compared to readmission (21.1% incidence) across all models and human evaluators. This counterintuitive finding, despite the lower incidence of mortality, may reflect a higher signal-to-noise ratio in the text related to mortality outcomes and warrants further exploration. It may be that the AI models and humans were able to associate particularly severe medical problems (e.g. end stage cancer) with likely mortality. However, readmission likelihood may vary with social factors such as availability of a caregiver at home, transportation factors, or other social determinants of health, which are less likely to be found in a discharge summary [10].

### Limitations

Several limitations of this study warrant discussion. This study was limited to open-source LLMs without fine tuning and excluded larger commercial models from providers like OpenAI, Google, and Anthropic, which may have achieved better performance.

Second, the dataset used, while extensive, is limited to a single institution, and might be insufficient to fully capture the variability in discharge summaries. A larger and more diverse dataset could potentially improve model robustness and generalizability.

Lastly, the physician evaluation involved only 100 discharge summaries, a significantly smaller sample than the 3,987 cases evaluated by ML models. A larger and more representative sample size for human evaluators would provide a stronger benchmark for comparison.

We encourage future studies focused on the evaluation of AI applications in healthcare. In particular, there is substantial experimentation in the fine-tuning of LLMs, which could potentially enhance their predictive performance. For example, researchers at New York University have developed NYUTron, a language model trained specifically using clinical care documents and have demonstrated promising performance in specific prediction tasks [11]. Future research should also evaluate commercially available LLMs, such as those from OpenAI, Anthropic, and Google, alongside other open-source LLMs, to determine their utility for these prediction tasks as well as combination software solutions extracting discrete data from other parts of the electronic health record in combination with LLM assessment of discharge summaries.

## Conclusion

Predicting 30-day mortality and readmission from discharge summaries remains a challenging endeavor. Despite leveraging a range of approaches, including machine learning models, COTS large language models (LLMs), and physician evaluations, none met our predefined clinical-grade reliability threshold of a Matthews Correlation Coefficient (MCC)>0.8. This limitation may be attributed to the low signal-to-noise ratio within discharge summaries for these predictive tasks or our current inability to extract this signal effectively. It is likely that additional data, not consistently found in the discharge summary, may be needed to generate reliable predictions for mortality and readmission. This study underscores the inherent complexity of predicting post-discharge outcomes. It highlights the importance of measuring AI performance with a robust metric and experimentation with diverse AI models.

## Data Availability

All data produced in the present work are contained in the manuscript

https://physionet.org/content/mimiciv/3.1/

**Approved for Public Release; Distribution Unlimited. Public Release Case Number 25-0765. ©2025 The MITRE Corporation. ALL RIGHTS RESERVED**

